# Oral Vitamin D supplementation induces transcriptomic changes in rectal mucosa that are consistent with anti-tumour effects

**DOI:** 10.1101/2021.05.04.21255629

**Authors:** P.G. Vaughan-Shaw, G. Grimes, JP Blackmur, M. Timofeeva, M Walker, L.Y Ooi, Victoria Svinti, Kevin Donnelly, FVN Din, S.M. Farrington, M.G. Dunlop

## Abstract

**Background:** Risk for several common cancers is influenced by the transcriptomic landscape of the respective tissue-of-origin. Vitamin D influences *in-vitro* gene expression and cancer cell growth. We sought to determine whether oral vitamin D induces beneficial gene expression effects in human rectal epithelium and identify biomarkers of response.

**Methods:** Blood and rectal mucosa was sampled from 191 human subjects and mucosa gene expression (HT12) correlated with plasma vitamin D (25-OHD) to identify differentially expressed genes. Fifty subjects were then administered 3200IU/day oral vitamin D3 and matched blood/mucosa resampled after 12 weeks’. Transcriptomic changes (HT12/RNAseq) after supplementation were tested against the prioritised genes for gene-set and GO-process enrichment. To identify blood biomarkers of mucosal response, we derived receiver-operator curves and C-statistic (AUC) and tested biomarker reproducibility in an independent Supplementation Trial (BEST-D).

**Results:** 629 genes were associated with 25-OHD level (P<0.01), highlighting 453 GO-term processes (FDR<0.05). In the whole intervention cohort, vitamin D supplementation enriched the prioritised mucosal gene-set (upregulated gene-set P<1.0E-07; downregulated gene-set P<2.6E-05) and corresponding GO terms (P=2.90E-02), highlighting gene expression patterns consistent with anti-tumour effects. However, only 9 individual participants (18%) showed a significant response (NM gene-set enrichment P<0.001) to supplementation. Expression changes in *HIPK2* and *PPP1CC* expression served as blood biomarkers of mucosal transcriptomic response (AUC=0.84 [95%CI:0.66-1.00]), and replicated in BEST-D trial subjects (*HIPK2* AUC=0.83 [95%CI:0.77-0.89]; *PPP1CC* AUC=0.91 [95%CI:0.86-0.95]).

**Conclusions:** Higher plasma 25-OHD correlates with rectal mucosa gene expression patterns consistent with anti-tumour effects and this beneficial signature is induced by short-term vitamin D supplementation. Heterogenous gene expression responses to vitamin D may limit the ability of randomised trials to identify beneficial effects of supplementation on CRC risk. However, in the current study blood expression changes in *HIPK2* and *PPP1CC* identify those participants with significant anti-tumor transcriptomic responses to supplementation in the rectum. These data provide compelling rationale for a trial of vitamin D and CRC prevention using easily assayed blood gene expression signatures as intermediate biomarkers of response.

## INTRODUCTION

Vitamin D deficiency is associated with risk of several common cancers, the strongest evidence supporting a link between vitamin D and colorectal cancer [1, 2]. However, a causal association has yet to be convincingly demonstrated, because the available observational evidence may be participant to several potential confounders. Environmental risk factors associated with CRC also associated with vitamin D status (i.e. co-causality; e.g., physical activity), while CRC or its treatment may itself lower plasma vitamin D levels (i.e. reverse causation). However, a recent randomised-control trial (RCT) reported an association between supplementation, vitamin D receptor genotype and risk of colorectal adenoma, supporting the premise that the beneficial effect may be causal [3]. Meanwhile, vitamin D-related genetic variation has been shown to influence the association between 25-OHD level and CRC survival [4-6], with a recent meta-analysis of RCT data strongly supporting a causal effect for vitamin D supplementation on CRC mortality [7, 8].

Differences in gene expression have been reported in CRC and adenoma tissue relative to normal colorectal tissue [9-12], with genes involved in metabolism, transcription and translation and cellular processes commonly altered [13]. Recent transcriptome wide association studies confirm importance of gene expression in carcinogenesis [14, 15]. Vitamin D broadly influences gene expression through activation of the ligand-activated transcription factor *VDR*, which has been shown to influence cancer cell growth *in vitro* [16]. Therefore, investigation of gene expression in the colorectum in the context of vitamin D status or supplementation may provide fresh insight into mechanisms underlying the relationship between CRC and vitamin D. Recent evidence suggests one mechanism may be that 1,25-dihydroxyvitamin D3 modulates immune and inflammatory pathway genes in large bowel epithelium [17]. However, differential expression in response to high dose 1,25-dihydroxyvitamin D3 may not accurately reflect the relationship between vitamin D status and gene expression at normal or low vitamin D levels, or in response to regular vitamin D3, the most commonly used vitamin D supplement.

We investigated whether circulating vitamin D concentration is associated with differential gene expression in rectal normal mucosa using a 2-Phase approach with validation of putative biomarkers in an independent study dataset. We directly assayed total 25-OHD, which reflects both dietary intake and skin synthesis of vitamin D [18, 19] and investigated its relationship with gene expression in normal mucosa and blood, assessed by microarray. In the Phase 1 correlative study we sought to identify a prioritised list of differentially expressed genes associated with 25-OHD level. In Phase 2, we conducted a study in human volunteers who were supplemented with oral vitamin D to determine whether the corresponding transcriptomic response was induced *in vivo*. Using blood peripheral blood mononuclear cells (PBMC) transcriptomic analysis we also identified potential blood biomarkers that indirectly indicate a beneficial response in the host rectal mucosa.

## METHODS

### Study Population

Participants recruited to Phase 1 of the Scottish Vitamin D study (SCOVIDS) (n=191) underwent sampling of blood and normal rectal mucosa by rigid sigmoidoscopic biopsy. RNA was extracted for gene expression analysis from matched contemporaneous NM and peripheral blood mononuclear cells (PBMCs). Plasma was collected at the same time for vitamin D analysis and DNA was extracted from whole blood for genotyping.

All eligible participants from Phase 1 were invited to proceed to Phase 2 which was an intervention study. Five of the 50 recruited participants to Phase 2 underwent interval sampling *before* starting supplementation to assess for longitudinal changes in expression before treatment. All Phase 2 participants were then administered vitamin D supplementation and underwent repeat NM, PBMC and 25-OHD sampling after 12 weeks’ 3200IU/ day cholecalciferol (Fultium-D3) supplementation (Figure 1). Concordance with the treatment protocol was assessed through a dose diary and pharmacy log of unused tablets (compliance of 98% of total doses taken achieved). Demographic and clinical data were prospectively collected from patient case notes.

**Figure 1.**
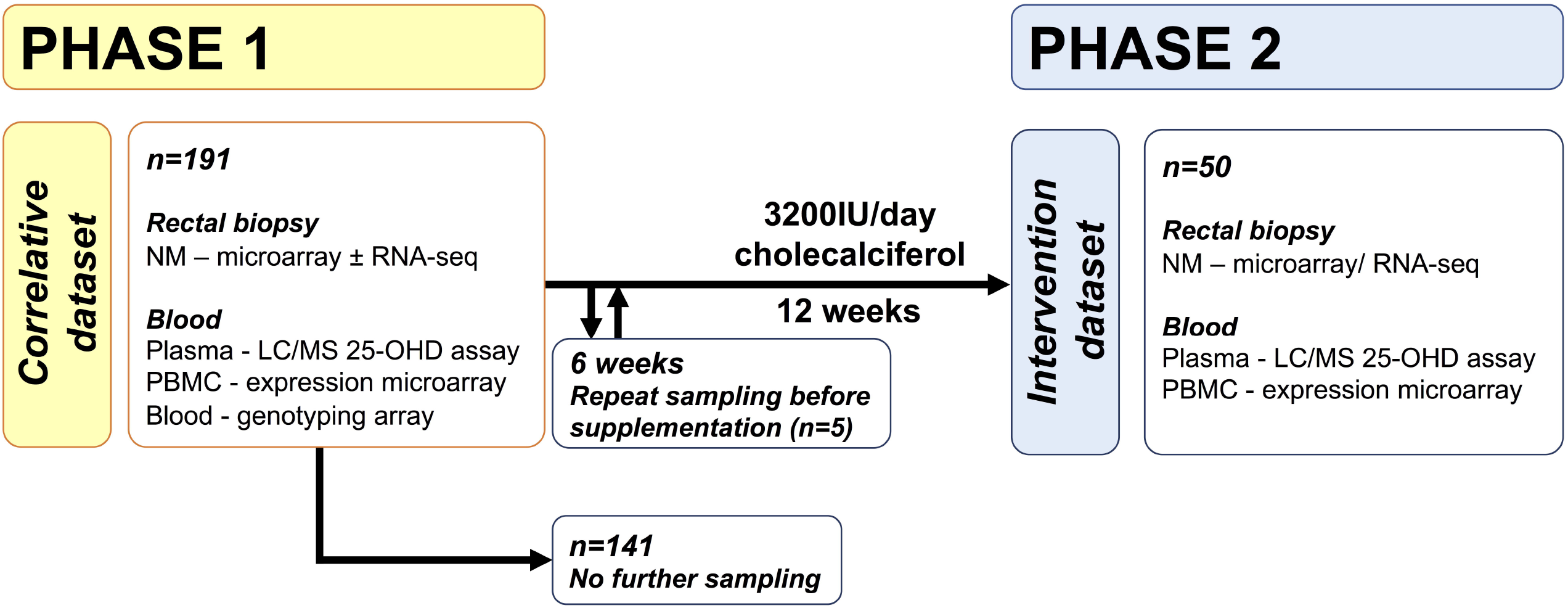
Summary of SCOVIDS study protocol. An unselected subset of Phase 1 participants (i.e. selection not based on 25-OHD or baseline gene expression) proceeded to Phase 2 and were given 3200IU cholecalciferol per day. Of these 5 participants were sampled 6 weeks after initial sampling and without supplementation to provide a control dataset, after which they proceeded to 12 week’s supplementation and final sampling.

### Sample size considerations

There were no available published data on which to base investigation of the sample size required to determine an association between vitamin D status and global gene expression in normal mucosa. Thus, a formal sample size estimation was not possible.

### Blood and mucosa sampling

Participants were sampled in outpatient clinic or during minor surgical procedures. No participant received cleansing oral mechanical bowel preparation. Blood was sampled by standard venepuncture of a peripheral arm vein, with plasma and PBMCs extracted (Supplementary Methods). A separate blood sample was taken to allow extraction of blood leukocyte DNA and genotyping of relevant vitamin D receptor and pathway SNPs. Normal rectal mucosa (NM) was sampled at the same time via rigid sigmoidoscopic rectal biopsy. NM and PBMC samples were immediately placed in RNAlater and kept immersed for 24-72 hours prior to RNA extraction or storage at -80□C.

### Plasma vitamin D assay

All plasma samples were measured to a standard, validated and published protocol by a single laboratory[20]. Total 25-OHD was measured by liquid chromatography tandem mass spectrometry.

### Assessment of gene expression

RNA was extracted and purified from NM and blood PBMC using a proprietary RNA extraction kit. Gene expression profiling was undertaken using the Illumina HumanHT-12v4.0 Expression BeadChip Arrays and IScan NO660 scanner, providing coverage of 47,231 transcripts and >31,000 annotated genes. For RNA sequencing, whole-genome transcriptomic patterns were analysed on total RNA from selected normal mucosa samples extracted as described above. RNA was sequenced on the Illumina HiSeq 2500 platform in “rapid mode” with 150bp paired-end reads in a single batch. Transcript indexing and quantification from RNA-seq reads was performed using Salmon v1.1.0[21].

### Statistical analysis

All statistical analysis was undertaken in R[22]. In Phase 1, linear regression modelling was used to test association between 25-OHD level and NM gene expression, adjusting for age, gender, CRC status and anaesthetic status (i.e. sampled under general anaesthetic). Genes associated with 25-OHD level at significance level P<0.01 were termed the ‘*candidate gene-set’* and taken forward for testing in the intervention dataset (Phase 2). In Phase 2, differences in 25-OHD level before and after vitamin D supplementation were investigated using paired Wilcoxon rank-sum test and differential gene expression analysis in response to vitamin D supplementation performed using the *lmFit* and *eBayes* functions within the ‘limma’ package [23] producing the intervention (Phase 2) dataset. Ranked lists of differentially expressed genes were assessed for functional relevance using the ‘GOrilla’, Gene Ontology enRIchment anaLysis and visuaLizAtion tool [24]. Process ontologies were investigated using gene lists ranked by coefficient (Phase 1) or fold-change (Phase 2).

We tested the Phase 2 dataset (i.e. response to supplementation) for enrichment of the Phase 1 candidate gene-set and top-ranked GO terms. Directional gene-set testing was performed in R, using the gene-setTest function in the ‘limma’ package [25]. We performed technical replication by performing gene-set enrichment testing on differential expression data derived from RNA-seq analysis of the same NM samples from the intervention cohort.

To identify biomarkers of response, we performed participant-level gene-set enrichment testing with a ‘response’ to supplementation defined as enrichment (P<0.001 given n=50 subjects) of the candidate gene-set after supplementation. Then, differentially expressed genes in the *blood* between those with/ without rectal NM response were tested for enrichment of the candidate gene-set. Logistic regression testing sought to identify potential blood biomarkers of response and utility of blood biomarkers was calculated using receiver operator curves and C statistic. Finally, we sought to validate putative biomarkers of response in an independent blood gene expression dataset derived from the ‘Biochemical Efficacy and Safety Trial of Vitamin D’ (BEST-D) study[26] (https://www.ebi.ac.uk/arrayexpress/files/E-MTAB-6246/).

## RESULTS

### Mucosal gene expression signature associated with higher 25-OHD level consistent with anti-tumor effects

In the Phase 1, 191 participants underwent rectal mucosal biopsy and blood sampling (Table 1). 25-OHD was nominally associated with expression of 629 probes (P<0.01), termed the ‘candidate gene-set’ (***‘Gene-set discovery’*** Figure 2).

**Table 1.**
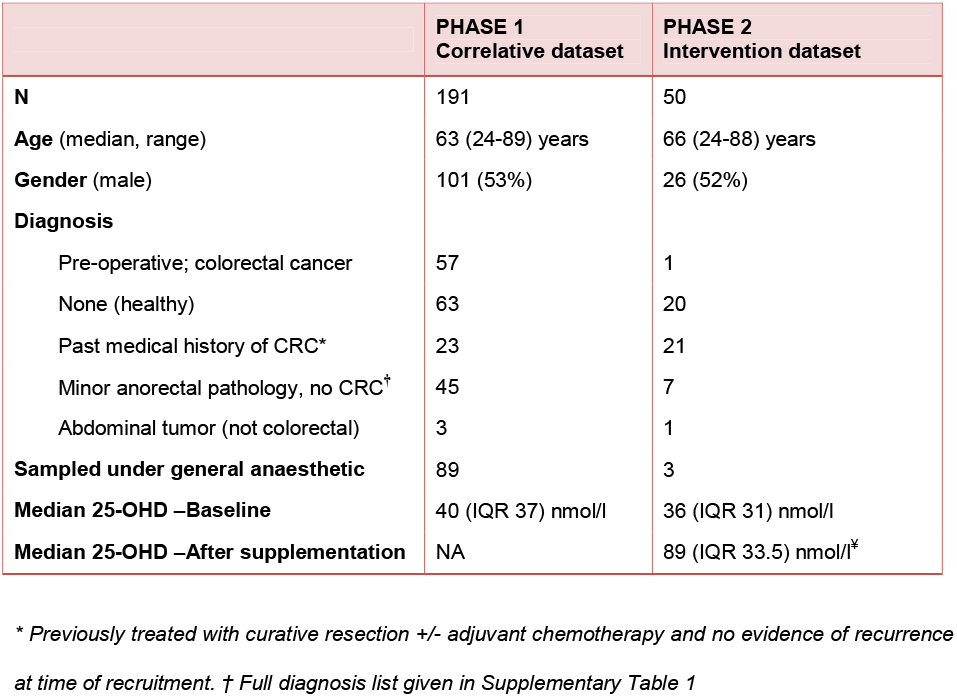
Baseline characteristics, sampling variables and vitamin D status in included participants.

**Figure 2.**
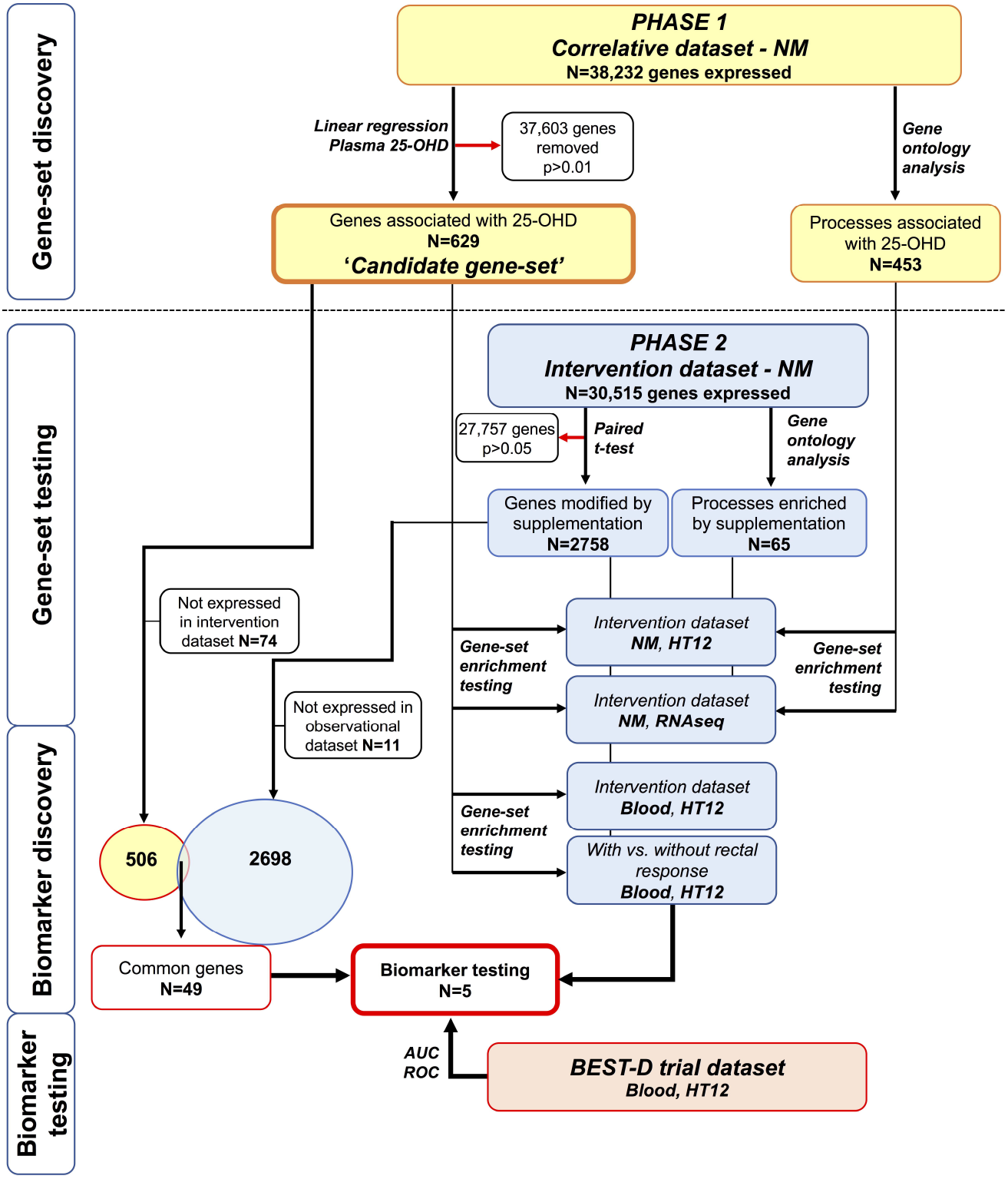
Analysis flowchart and gene-set selection for biomarker assessment.

No individual probe was significantly associated with 25-OHD after adjustment for genome-wide multiple testing (Supplementary Table 2), yet the top three hits have previous reported association with colorectal tumorigenesis *CNN1*[27], *COX7A1*[28], *PIP5K1C*[29]. Gene ontology analysis demonstrated significant enrichment of 453 processes (Supplementary Table 3) with many highly relevant to carcinogenesis e.g. ‘*regulation of cell migration’* (FDR=7.55E-08), ‘*regulation of programmed cell death’* (FDR=5.38E-03), and ‘*regulation of cell differentiation’* (FDR=2.55E-05). Several genes from the candidate gene-set with higher expression associated with higher 25-OHD are included in enriched GO ontology terms relevant to carcinogenesis, and have reported tumor suppressor activity (e.g. *FOXOs, CAV1, LRP1*, Supplementary Tables 4 and 5). This suggests that the NM gene expression signature, i.e. Phase 1 ‘candidate gene-set’, associated with higher 25-OHD level is consistent with anti-tumor effects.

### Oral vitamin D supplementation enriches anti-tumor expression signature in normal rectal mucosa

In Phase 2, 50 participants were administered vitamin D supplementation and underwent repeat sampling after 12-weeks’. Post-hoc analysis revealed age, gender and baseline 25-OHD to be similar between Phase 1/2 participants (P>0.05). Supplementation induced an increase in plasma 25-OHD after 12-weeks (median plasma 25-OHD before/after supplementation was 36nmol/l, 89nmol/l; P= 2.5E-09, Supplementary Table 6).

No individual gene from the candidate gene-set showed significant differential expression after adjustment for multiple testing (Supplementary Table 7). However, testing of the Phase 1 candidate gene-set showed significant enrichment after supplementation (upregulated gene-set P<1.0E-07; downregulated gene-set 2.8E-05, see **‘Gene-set testing’** Figure 2, Table 2, Supplementary Figure 1), confirmed in RNA-seq data.

**Table 2.**
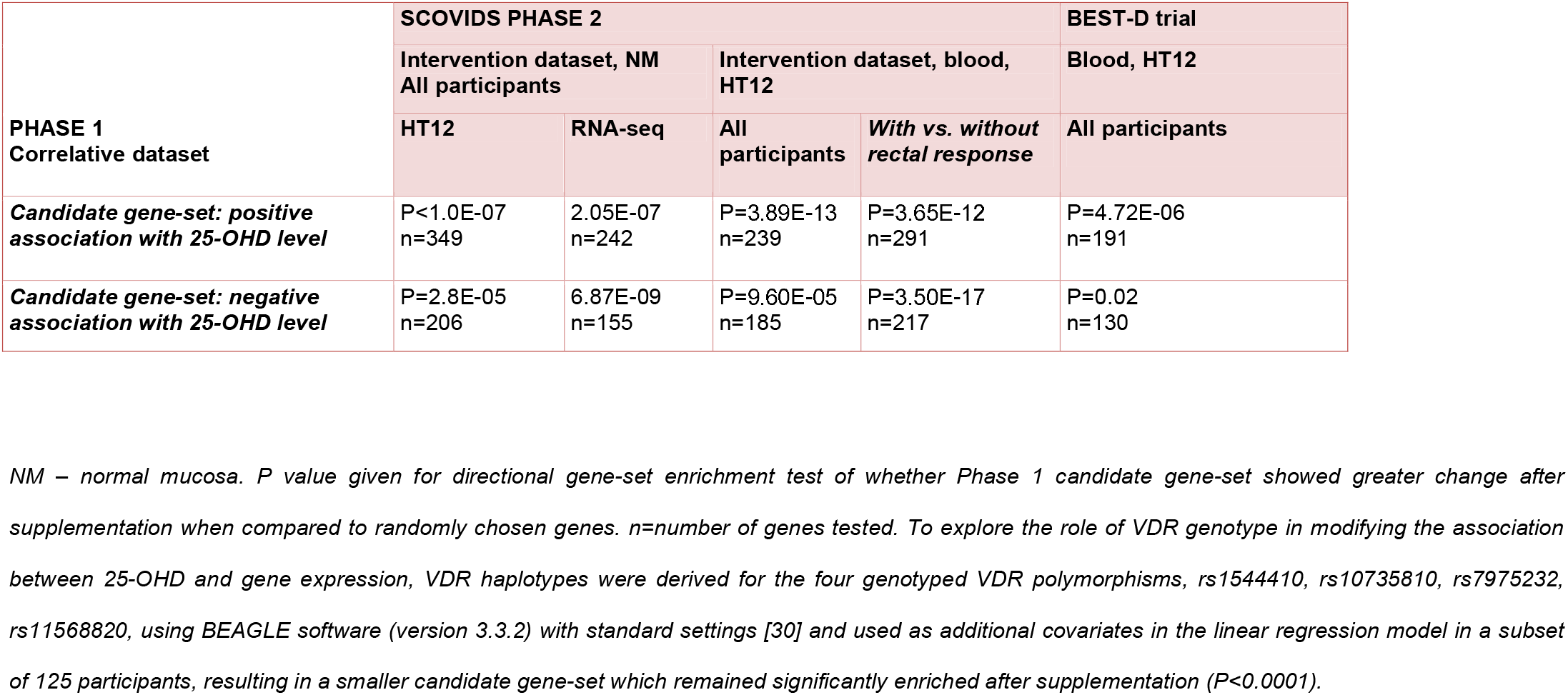
Gene-set testing for enrichment of the candidate gene-set from Phase 1 after supplementation in Phase 2 and the BEST-D study.

Enrichment of the candidate gene-set was not associated with 25-OHD response to supplementation, with those with the lowest 25-OHD FCs still showing gene-set enrichment (Supplementary Figure 1). Meanwhile, there was no enrichment of the candidate gene-set in interval NM samples taken *before* commencement of supplementation indicating it is a treatment effect (median interval 8 weeks, Supplementary Figure 1).

Of the 629 candidate genes associated with circulating vitamin D in the Phase 1 gene-set, fifty-five had nominally significant expression change after supplementation (P<0.05). Concordance in direction of effect between the coefficient of association with plasma 25-OHD level and expression change after supplementation was observed in 49 of these (R0.93, P<2.2E-16, Supplementary Table 7), with these genes taken forward for biomarker discovery (see ‘**Biomarker discovery’** section Figure 2).

### GO term enrichment indicates modulation of anti-tumor effects in normal mucosa by supplementation

Functional annotation of the intervention dataset gene list identified 65 significantly enriched pathways after supplementation, with many terms relevant to carcinogenesis including *‘regulation of programmed cell death* (FDR=9.66E-03) and *‘regulation of cell migration’* (FDR=7.83E-03) (Supplementary Table 8). Taken together, genes in the top 50 GO terms from Phase 1 were significantly enriched after supplementation (P=2.90E-02). Common processes across both the Phase 1 and Phase 2 datasets included terms relevant to carcinogenesis including *‘regulation of programmed cell death’, ‘regulation of cell migration’* (Supplementary Figure 2), demonstrating that biologically relevant patterns of gene expression changes associated with higher 25-OHD level and consistent with anti-tumor effects could be imparted by oral supplementation.

### Blood expression biomarkers identify participants with gene expression response to supplementation

We identified 9 individual participants (18%) with a significant response (i.e. candidate gene-set enrichment in NM P<0.001) to supplementation. Of the top 50 ranked GO terms from Phase 1, 43 were enriched in these 9 participants after supplementation indicating a biologically relevant NM gene expression response to supplementation, which was absent in the remaining participants (Supplementary Table 9). NM gene expression response was associated with an increased allele risk score of the four functionally relevant *VDR* SNPs (p=0.006), but not with *VDR* gene expression or increased 25-OHD fold-change (Supplementary Table 10).

Changes in PBMC gene expression after supplementation reflected those in the rectum, with the Phase 1 candidate gene-set significantly enriched in blood after supplementation (Table 2). Moderate correlation between blood and rectum fold-change in the 49 genes taken forward for biomarker discovery was seen (R=0.64, P=5.63E-06). When we compared PBMC gene expression after supplementation between those participants *with* and *without* a rectal mucosal gene expression response, the differentially expressed genes in PBMCs were enriched for the candidate gene-set, indicating potential blood biomarkers of mucosal response (Table 2, see ‘**Biomarker discovery’** section Figure 2). Five genes identified from the Phase 1 which were both differentially expressed in NM after supplementation and also differentially expressed in blood between participants *with* and *without* a rectal response to supplementation (*SMEK2, HIPK2, PPP1C, DDR1 and SNX21*), indicating biomarker potential (Table 3). When the genes were combined, a blood expression signature based on the best derived cut-off showed strong utility in predicting NM response (AUC=0.99, 95%CI: 0.97-1.00, Supplementary Table 10, Supplementary Figure 4).

**Table 3.**
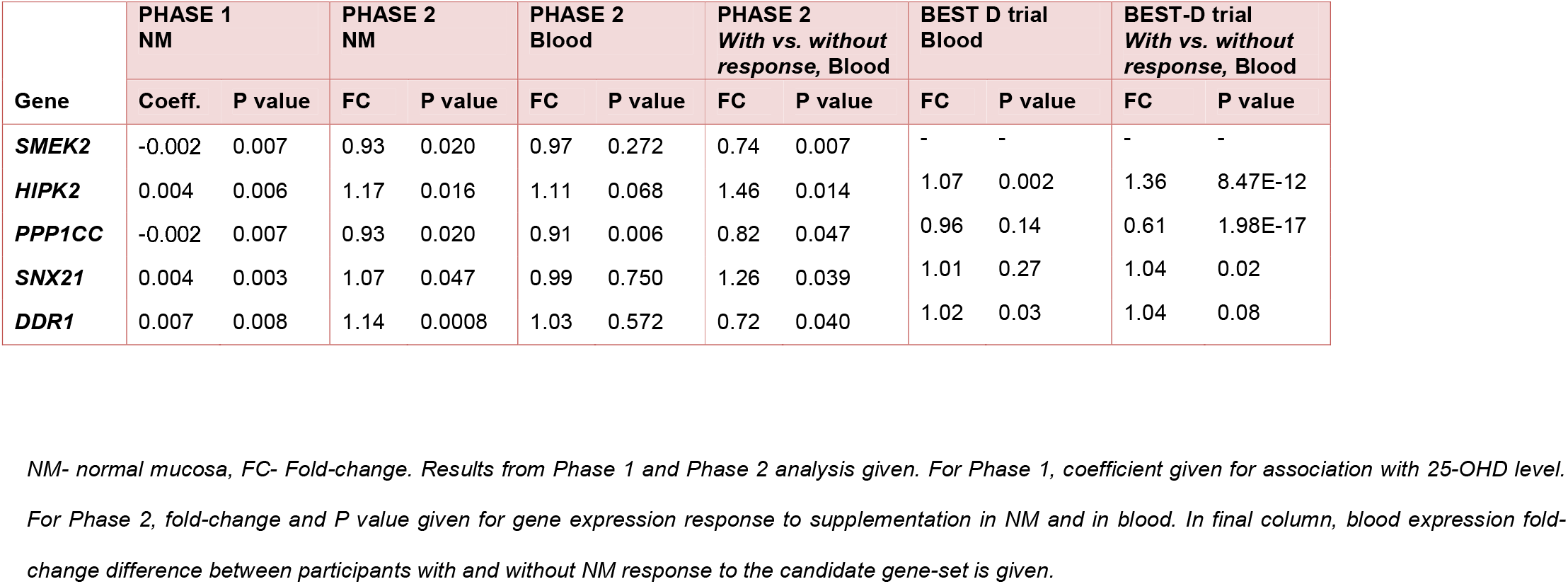
Genes prioritised from Phase 1 correlative dataset modified by supplementation in NM with evidence of potential biomarker utility in PHASE 2 and BEST-D trial.

We then explored the value of these same genes as blood biomarkers of *blood* response to supplementation. The *HIPK2* and *PPP1CC* genes (Table 3, Figure 3) demonstrated the best predictive utility in identifying participants with response in both rectum (AUC=0.84, 95%CI: 0.66-1.00) and blood (AUC=0.87, 95%CI: 0.71-1.00, Supplementary Table 11).

**Figure 3.**
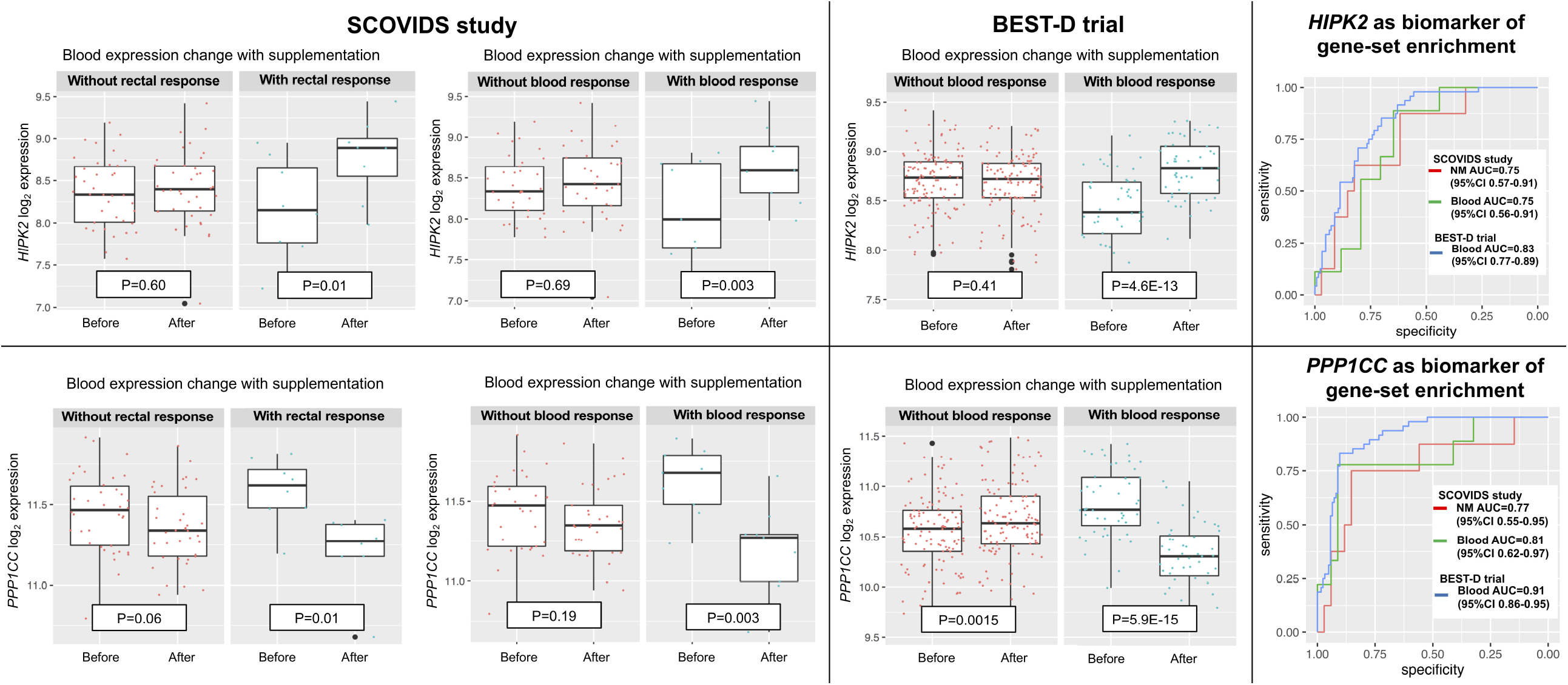
Blood *HIPK2* and *PPP1CC* expression before and after supplementation in SCOVIDS and BEST-D trial with ROC of biomarker utility. Response defined as participant level gene-set enrichment to Phase 1 candidate gene-set from our SCOVIDS study after HIPK2, PPP1CC, SMEK2, DDR1, SNX21 excluded.

### *HIPK2* and *PPP1CC* are independent biomarkers of expression response to supplementation in the BEST-D trial expression dataset

In the BEST-D trial, 48/172 (28%) participants showed significant enrichment of our Phase 1 candidate gene-set. *HIPK2* expression increased with supplementation in the BEST-D trial (FC=1.07, p=0.002), with the increase greatest in those with a blood response to supplementation as defined by our candidate gene-set (P=4.6E-13, Figure 3) and a 1.36 fold-difference in expression change after supplementation between those with/ without a response to our candidate gene-set (P=8.48E-12, Table 3). In the current intervention study (SCOVIDS), we observed an average *HIPK2* FC=1.11 in blood, with 21 (47%) participants showing a FC>1.19, the optimum threshold determined by AUC calculations. In the BEST-D study, 63 (37%) participants taking oral vitamin D supplementation showed a FC>1.19 response, with this signature more prevalent in those taking 4000IU per day (42%), suggesting a dose-response. *HIPK2* expression change showed utility in identifying those with a response to our candidate gene-set, AUC=0.83 (95%CI:0.77-0.89, Figure 3) and when the threshold from our study was used, *HIPK2* FC>1.19 AUC=0.74 (95%CI: 0.66-0.81, Supplementary Table 10).

PPP1CC expression showed non-significant decrease after supplementation in the BEST-D cohort overall, but when the participants were stratified by blood response, *PPP1CC* was seen to increase in those without blood response (P=0.0015, Figure 3) and markedly decreased in those with a blood response (P= 5.9E-15). There was a 0.61 fold-difference in *PPP1CC* expression change between those with and without a response to our candidate gene-set (P=1.98E-17, Table 3). A total of 47 (27%) participants had *PPP1CC* FC<0.76 after supplementation, closely reflecting the 11 (26%) participants in the current intervention study. Crucially, *PPP1CC* expression change after supplementation showed utility as a biomarker of blood response to our candidate gene-set, AUC=0.91 (95%CI=0.86-0.95, Figure 3) and when using the threshold from the SCOVIDs study, *PPP1CC* FC<0.76 AUC=0.83 (95%CI:0.76-0.89, Supplementary Table 10).

## DISCUSSION

This study reveals demonstrable differences in gene expression patterns in normal rectal mucosa correlated with plasma 25-OHD level. These differences are consistent with beneficial effects on processes relevant to colorectal carcinogenesis. Furthermore, we show that oral supplementation with vitamin D induces changes in the prioritised gene list. This indicates that the beneficial expression “signature” is not static, but rather can be modified by oral vitamin D supplementation, at least within the timescale tested here. Although we were not able to directly test cancer endpoints, there is considerable published evidence supporting the premise that enrichment of this favourable gene-set imparts anti-tumor effects.

Homeodomain-interacting protein kinase 2 (*HIPK2*) is a known tumor suppressor gene [31] and the Protein Phosphatase 1 Catalytic Subunit Gamma gene (*PPP1CC*), a published molecular marker of CRC [32]. We found expression changes in these genes in blood to have predictive value in reflecting rectal mucosa response to supplementation. Hence, these may have utility as blood biomarkers of a beneficial epithelial response to supplementation. In addition, the effect on gene expression in blood PBMCs appears robust, since we replicated the effect in a large, independent, expression dataset, namely the BEST-D trial in which subjects were administered oral vitamin D supplementation (2000/4000IU 12 months).

We devised a 2-Phase *in vivo* approach, firstly to identify differentially expressed genes in the rectal epithelium associated with plasma 25-OHD and determine the GO terms and processes linked to that prioritised gene list. However, our ultimate aim was to establish whether these transcriptomic responses could be recapitulated by oral vitamin D supplementation, thereby demonstrating a modifiable transcriptomic landscape. Many of the top-ranked genes associated with higher 25-OHD level have links with CRC, for instance *CNN1* [27], *COX7A1* [28], PEG3 [33], *PIP5K1C* [29], *TAGLN* [34], *DAAM2* [35]. Furthermore, we highlight a number of genes within processes relevant to tumorigenesis which are associated with 25-OHD level and influenced by supplementation. The directions of effect of these genes were consistent with tumor suppressor activity. Enrichment of pathways involved in cell migration and cell death validate published *in vitro* data which demonstrate vitamin D-induced growth arrest and apoptosis of CRC cells lines, modulation of the *Wnt* signalling pathway, DNA repair and immunomodulation [16]. Published clinical data also corroborate our current findings: Protiva *et al*., reported upregulation of genes involved in inflammation, immune response, extracellular matrix, and cell adhesion in response to 1,25(OH)2D3 [17], while Fedirko *et al*., reported a reduction in tumor promoting inflammation biomarkers, decreased oxidative DNA damage, increased cell differentiation and apoptosis and modification of the APC/β-catenin pathway in the normal human colorectal epithelium [36]. Taken together, these data suggest possible mechanisms underlying the widely reported link between vitamin D deficiency and increased CRC risk [1, 2]. It also might explain the recently reported beneficial impact of supplementation on CRC survival outcomes [8].

Despite compelling published observational and pre-clinical data, the link between vitamin D and risk of cancer and several other traits remains controversial. Indeed, several large intervention trials have shown no benefit on cancer endpoints (VITAL Trial [37], Vitamin D Assessment (ViDA) study [38, 39] and Baron *et al*. [40]). However, participants in these trials were predominantly sufficient for vitamin D at the trial outset, thereby potentially blunting beneficial effects [41]. We have previously rehearsed potential reasons why previous study designs might have failed to detect real effects [41]. To counter potential confounding effects, we conducted a Mendelian randomisation study but this also did not demonstrate a beneficial effect of circulating vitamin D on CRC risk. However, available genetic instrumental variables are weak and explain only a small portion of variance of 25OHD levels [42, 43].

In this study, 18% of participants receiving vitamin D supplementation exhibited a response in the colorectal epithelium (the putative target tissue). If our hypothesis holds that expression changes translate to cancer endpoints, this low response rate would adversely impact on statistical power of trials conducted to date which have tested the effect of vitamin D supplementation on clinical endpoints. Such trials routinely perform subgroup analyses based on change in circulating 25-OHD level, yet the current study reveals poor correlation between plasma level and mucosal gene expression changes, suggesting non-linear responses to vitamin D may introduce further heterogeneity to clinical endpoints.

We have identified blood biomarkers that reliably identify participants who respond to vitamin D supplementation by inducing gene expression changes in the target tissue. The value of these biomarkers is replicated in a larger independent expression dataset. Further work is required to assess the utility of respective blood protein assays (e.g. ELISA) and reproducibility of these blood biomarkers in identifying mucosal response across a larger cohort. Nevertheless, these exciting and novel findings provide rationale for a trial of vitamin D and CRC prevention using easily assayed blood gene expression signatures as intermediate biomarkers of response.

Whilst the 2-Phase design of an intervention study informed by our correlative dataset has many positive attributes, the study has a number of limitations. First, the low median level of 25-OHD and narrow positively skewed distribution of 25-OHD in the Phase 1 cohort may have masked some true associations between vitamin D level and gene expression. Failure to identify individual gene significance after adjustment for genome-wide multiple testing may also indicate inadequate sample size, physiological autoregulation maintaining constant gene expression despite differences in circulating 25-OHD, or differences between plasma and rectal mucosa concentrations of 25-OHD or 1,25-OHD [44].

This intervention study is larger than many published studies of gene expression and vitamin D supplementation [17, 45-48], yet may still have limited power to achieve individual gene significance. Phase 2 participants were recruited as a subset of the Phase 1 cohort, which may influence gene-set enrichment test results. However, we did not select those who received supplementation based on 25-OHD level or baseline gene expression, which could have led to overfitting of the data, but instead took an unselected group. If anything, this approach could blunt the observed effect of supplementation, as mucosal response to supplementation may be capped in those with specific 25-OHD or favorable patterns of gene expression at baseline. Despite this we observed significant enrichment of the candidate gene-set derived in Phase 1 in those receiving supplementation. Sampling of rectal mucosa but not colonic mucosa avoided the use of cleansing bowel laxatives which may influence gene expression [49], yet limits the generalisability of our findings to more proximal colonic mucosa. Finally, sampling after 12-weeks of supplementation may not adequately capture early or later gene expression changes yet more frequent or delayed sampling would provide additional practical and ethical challenges.

In conclusion, we report for the first time patterns of gene expression and functional pathways in the normal rectal mucosa that are associated with circulating plasma vitamin D level. Oral vitamin D supplementation induces transcriptomic changes consistent with beneficial anti-tumor effects. Blood leukocyte expression of *HIPK2* and *PPP1CC* predicted well those participants with the greatest expression response following supplementation. Whilst further replication in a separate cohort is desirable, these data provide compelling rationale for a trial of vitamin D and CRC prevention using easily assayed blood gene expression signatures as intermediate biomarkers of response.

## Data Availability

Available at https://www.ncbi.nlm.nih.gov/geo/ GEO ID GSE157982. Full phenotypic data available from the corresponding author on reasonable request.

## Glossary

25-OHD: 25-hydroxyvitamin D
AUC: Area under curve
BEST-D: Biochemical Efficacy and Safety Trial of Vitamin D trial
CRC: colorectal cancer
DNA: Deoxyribonucleic acid
FC: fold-change
GO: Gene ontology
NM: normal mucosa
PBMC: peripheral blood mononuclear cells
RCT: randomised control trial
RNA: ribonucleic acid
SCOVIDS: Scottish Vitamin D study
SNP: single nucleotide polymorphism
VDR: vitamin D receptor gene

## ADDITIONAL FILES

### Supplementary Methods (docx)

Supplementary methods to be read as an adjunct to the main methods section in the manuscript.

### Supplementary Tables (docx)

Supplementary Tables to be read as an adjunct to the main methods section in the manuscript.

### Protocol (doc)

Full protocol for SCOVIDS study

## Acknowledgements

We acknowledge statistical advice from Dr Catalina Vallejos, Biomedical Data Science research group, University of Edinburgh. We acknowledge the excellent technical support from Stuart Reid. We are grateful to Donna Markie, and all those who continue to contribute to recruitment, data collection, and data curation for the Scottish Vitamin D Study. We acknowledge the expert support on sample preparation from the Genetics Core of the Edinburgh Wellcome Trust Clinical Research Facility in addition to the nursing and study facilities provided by the Clinical Research Facility.

## DECLARATIONS

### Ethical approval and consent

All participants provided informed written consent. The research was approved by the South East Scotland Research Ethics Committee 03 (11/SS/0109), South East Scotland Research Ethics Committee 01 (13/SS/0248) and Lothian National Health Service Research and Development office (2014/0058).

### Author contributions

Conceptualization, PGVS, MGD; Methodology, PGVS, LYO, GG, ET, FVND, SMF, MGD; Investigation, PGVS, LYO, JPB, GG, MT, MW, VS, KD; Writing Original Draft PVS, SMF, MGD; Writing – Review & Editing, JPB, MT, SMF, FVND, MGD; Funding Acquisition, MGD.; Resources, MGD; Supervision, SMF and MGD.

**Author names in bold designate shared co-senior authorship**

