## Supplementary Tables for "Oral Vitamin D supplementation induces transcriptomic changes in rectal mucosa that are consistent with anti-tumour effects"

Supplementary tables and figures

Supplementary Table 1 Full list of diagnoses in recruited participants

|  | PHASE 1  Correlative dataset | PHASE 2  Intervention dataset |
| --- | --- | --- |
| N | 191 | 50 |
| Diagnosis |  |  |
| Pre-operative; colorectal cancer | 57 | 1 |
| None (healthy) | 63 | 20 |
| Past medical history of CRC | 23 | 21 |
| Abdominal tumour (not colorectal) | 3 | 1 |
| Minor anorectal pathology, no CRC | 45 | 7 |
| Haemorrhoids | 20 | 1 |
| Fistula-in-ano | 7 | 2 |
| Anal intraepithelial neoplasia | 4 | 2 |
| Fissure-in-ano | 4 | - |
| Anal fibroepithelial polyp | 3 | - |
| Pilonidal disease | 3 | 1 |
| Anal warts | 1 | - |
| Anal skin tag | 2 | 1 |
| Hidradenitis suppurativa | 1 | - |

Supplementary Table 2 Top 50 ranked genes in PHASE 1

Linear regression modelling was applied to all probes on the HT12, adjusted by age, gender, CRC status and anaesthetic status. Top 50 hits ranked by p value listed

| Probe | *Gene* | Estimate | P value | Adjusted P value |
| --- | --- | --- | --- | --- |
| ILMN_1668514 | *PIP5K1C* | 0.005 | 2.69E-05 | 0.39 |
| ILMN_1810054 | *CNN1* | 0.015 | 3.67E-05 | 0.39 |
| ILMN_1662419 | *COX7A1* | 0.009 | 4.25E-05 | 0.39 |
| ILMN_2045994 | *SEPW1* | 0.007 | 4.41E-05 | 0.39 |
| ILMN_1798210 | *E2F7* | -0.003 | 7.38E-05 | 0.39 |
| ILMN_1761968 | *PPP1R14A* | 0.007 | 8.00E-05 | 0.39 |
| ILMN_1655557 | *INTS6* | -0.004 | 8.68E-05 | 0.39 |
| ILMN_1733851 | *DACT3* | 0.007 | 8.77E-05 | 0.39 |
| ILMN_1697200 | *MON2* | -0.003 | 1.36E-04 | 0.39 |
| ILMN_1805842 | *FHL1* | 0.007 | 1.37E-04 | 0.39 |
| ILMN_1752668 | *DAAM2* | 0.004 | 1.39E-04 | 0.39 |
| ILMN_1763524 | *LZTS2* | 0.005 | 1.46E-04 | 0.39 |
| ILMN_1781388 | *PGM5* | 0.010 | 1.48E-04 | 0.39 |
| ILMN_1709590 | *PGM5* | 0.010 | 1.61E-04 | 0.39 |
| ILMN_2389876 | *TGFB1I1* | 0.007 | 1.61E-04 | 0.39 |
| ILMN_1778668 | *TAGLN* | 0.009 | 1.84E-04 | 0.39 |
| ILMN_1712634 | *TIA1* | -0.004 | 1.87E-04 | 0.39 |
| ILMN_1680569 | *SSX5* | 0.001 | 1.88E-04 | 0.39 |
| ILMN_1771139 | *FBXO31* | 0.003 | 1.93E-04 | 0.39 |
| ILMN_2368834 | *MYH11* | 0.013 | 2.20E-04 | 0.42 |
| ILMN_1804955 | *CTSF* | 0.005 | 2.31E-04 | 0.42 |
| ILMN_2271149 | *PGM5* | 0.009 | 2.82E-04 | 0.45 |
| ILMN_1653200 | *SLC22A17* | 0.004 | 2.94E-04 | 0.45 |
| ILMN_1807493 | *ACVRL1* | 0.007 | 3.17E-04 | 0.45 |
| ILMN_1798620 | *PQLC1* | 0.003 | 3.17E-04 | 0.45 |
| ILMN_1757604 | *TPM2* | 0.010 | 3.21E-04 | 0.45 |
| ILMN_1747244 | *CCNG2* | -0.005 | 3.32E-04 | 0.45 |
| ILMN_1791494 | *SYNPO2* | 0.008 | 3.60E-04 | 0.45 |
| ILMN_3244323 | *LOC148413* | 0.003 | 3.77E-04 | 0.45 |
| ILMN_1712075 | *SYNM* | 0.011 | 3.80E-04 | 0.45 |
| ILMN_1765409 | *STAM* | -0.004 | 3.99E-04 | 0.45 |
| ILMN_1808238 | *RBPMS2* | 0.010 | 4.04E-04 | 0.45 |
| ILMN_1667692 | *PTGIS* | 0.003 | 4.18E-04 | 0.45 |
| ILMN_1709818 | *LOC643721* | -0.001 | 4.29E-04 | 0.45 |
| ILMN_3255569 | *LOC100129303* | 0.002 | 4.40E-04 | 0.45 |
| ILMN_3236858 | *NYNRIN* | 0.007 | 4.46E-04 | 0.45 |
| ILMN_2374293 | *DYRK1A* | -0.004 | 4.80E-04 | 0.45 |
| ILMN_1720048 | *CCL2* | 0.009 | 4.94E-04 | 0.45 |
| ILMN_1660086 | *MYH11* | 0.014 | 4.97E-04 | 0.45 |
| ILMN_2388466 | *TIA1* | -0.004 | 5.27E-04 | 0.45 |
| ILMN_1687279 | *DHPS* | 0.003 | 5.30E-04 | 0.45 |
| ILMN_1798790 | *IL17RC* | 0.005 | 5.33E-04 | 0.45 |
| ILMN_1721283 | *HSPB6* | 0.008 | 5.36E-04 | 0.45 |
| ILMN_1880222 | *LOC731496* | 0.001 | 6.02E-04 | 0.45 |
| ILMN_3215381 | *LOC645175* | -0.004 | 6.03E-04 | 0.45 |
| ILMN_1675331 | *PEG3* | 0.002 | 6.14E-04 | 0.45 |
| ILMN_2219767 | *MYCN* | -0.004 | 6.17E-04 | 0.45 |
| ILMN_3308103 | *MIR1321* | 0.001 | 6.18E-04 | 0.45 |
| ILMN_1717636 | *RGMA* | 0.005 | 6.18E-04 | 0.45 |
| ILMN_1741389 | *B3GNT8* | 0.007 | 6.21E-04 | 0.45 |

Supplementary Table 3 Top 50 ranked GO terms enriched in PHASE 1

| GO term | Description | p-value | FDR q-value | Enrichment |
| --- | --- | --- | --- | --- |
| GO:0040011 | Locomotion | 2.46E-14 | 3.78E-10 | 1.85 |
| GO:0016477 | Cell migration | 9.25E-14 | 7.09E-10 | 2.01 |
| GO:0009653 | Anatomical structure morphogenesis | 1.64E-13 | 8.39E-10 | 1.92 |
| GO:0048870 | Cell motility | 1.74E-13 | 6.66E-10 | 1.88 |
| GO:0043062 | Extracellular structure organization | 2.69E-13 | 8.25E-10 | 2.32 |
| GO:0065008 | Regulation of biological quality | 4.92E-13 | 1.26E-09 | 1.38 |
| GO:0022603 | Regulation of anatomical structure morphogenesis | 5.12E-13 | 1.12E-09 | 1.77 |
| GO:0030198 | Extracellular matrix organization | 5.73E-13 | 1.10E-09 | 2.31 |
| GO:0006928 | Movement of cell or subcellular component | 6.05E-13 | 1.03E-09 | 1.68 |
| GO:0032502 | Developmental process | 8.34E-13 | 1.28E-09 | 1.33 |
| GO:0006936 | Muscle contraction | 2.29E-12 | 3.19E-09 | 31.78 |
| GO:0050793 | Regulation of developmental process | 6.52E-12 | 8.33E-09 | 1.45 |
| GO:0048646 | Anatomical structure formation involved in morphogenesis | 6.80E-12 | 8.01E-09 | 1.85 |
| GO:0048856 | Anatomical structure development | 9.07E-12 | 9.93E-09 | 1.4 |
| GO:0051128 | Regulation of cellular component organization | 1.45E-11 | 1.49E-08 | 1.49 |
| GO:0003012 | Muscle system process | 1.56E-11 | 1.50E-08 | 26.08 |
| GO:0022610 | Biological adhesion | 3.82E-11 | 3.44E-08 | 2.59 |
| GO:0097435 | Supramolecular fibre organization | 4.64E-11 | 3.95E-08 | 2.11 |
| GO:0007155 | Cell adhesion | 5.70E-11 | 4.60E-08 | 1.83 |
| GO:0051094 | Positive regulation of developmental process | 6.34E-11 | 4.86E-08 | 1.63 |
| GO:0010273 | Detoxification of copper ion | 7.74E-11 | 5.65E-08 | 35.17 |
| GO:0051239 | Regulation of multicellular organismal process | 8.42E-11 | 5.87E-08 | 1.38 |
| GO:0048518 | Positive regulation of biological process | 9.09E-11 | 6.05E-08 | 1.28 |
| GO:0030334 | Regulation of cell migration | 1.18E-10 | 7.55E-08 | 1.76 |
| GO:2000145 | Regulation of cell motility | 1.63E-10 | 1.00E-07 | 1.73 |
| GO:0061687 | Detoxification of inorganic compound | 1.84E-10 | 1.09E-07 | 32.65 |
| GO:0046688 | Response to copper ion | 2.12E-10 | 1.20E-07 | 17.26 |
| GO:0048583 | Regulation of response to stimulus | 2.46E-10 | 1.35E-07 | 1.31 |
| GO:0051270 | Regulation of cellular component movement | 2.55E-10 | 1.35E-07 | 1.7 |
| GO:0043408 | Regulation of MAPK cascade | 2.85E-10 | 1.46E-07 | 2.07 |
| GO:0048522 | Positive regulation of cellular process | 3.49E-10 | 1.72E-07 | 1.3 |
| GO:0040012 | Regulation of locomotion | 4.01E-10 | 1.92E-07 | 1.69 |
| GO:0040008 | Regulation of growth | 5.12E-10 | 2.38E-07 | 1.81 |
| GO:0007166 | Cell surface receptor signalling pathway | 5.14E-10 | 2.32E-07 | 1.4 |
| GO:0048523 | Negative regulation of cellular process | 6.11E-10 | 2.68E-07 | 1.25 |
| GO:0010035 | Response to inorganic substance | 9.66E-10 | 4.11E-07 | 2.17 |
| GO:0045597 | Positive regulation of cell differentiation | 1.83E-09 | 7.57E-07 | 1.71 |
| GO:0098754 | Detoxification | 2.51E-09 | 1.01E-06 | 8.09 |
| GO:0048519 | Negative regulation of biological process | 3.36E-09 | 1.32E-06 | 1.21 |
| GO:0023051 | Regulation of signalling | 3.85E-09 | 1.48E-06 | 1.32 |
| GO:0009966 | Regulation of signal transduction | 3.86E-09 | 1.44E-06 | 1.35 |
| GO:0051272 | Positive regulation of cellular component movement | 4.16E-09 | 1.52E-06 | 1.91 |
| GO:0030029 | Actin filament-based process | 4.19E-09 | 1.49E-06 | 2.12 |
| GO:0006882 | Cellular zinc ion homeostasis | 4.58E-09 | 1.60E-06 | 26.23 |
| GO:0048585 | Negative regulation of response to stimulus | 4.80E-09 | 1.63E-06 | 1.44 |
| GO:0042127 | Regulation of cell proliferation | 5.49E-09 | 1.83E-06 | 1.45 |
| GO:0042221 | Response to chemical | 5.67E-09 | 1.85E-06 | 1.43 |
| GO:2000026 | Regulation of multicellular organismal development | 6.16E-09 | 1.97E-06 | 1.43 |
| GO:0040017 | Positive regulation of locomotion | 6.44E-09 | 2.01E-06 | 1.89 |
| GO:0009887 | Animal organ morphogenesis | 7.07E-09 | 2.17E-06 | 2.39 |

Supplementary Table 4 Candidate genes positively associated with 25-OHD level in the GO term ‘*regulation of* *cell migration’* and published evidence of tumour suppressor/ biomarker activity

21 genes from the candidate gene-set were associated with GO term regulation of cell migration, with a total of 166 genes associated with this term.

| ILMN ID | Gene | Coefficient | P value | Reference |
| --- | --- | --- | --- | --- |
| ILMN_1680973 | *FOXF1* | 0.005 | 0.010 | [1] |
| ILMN_1712095 | *FOXO4* | 0.003 | 0.002 | [2] |
| ILMN_1789599 | *NBL1* | 0.008 | 0.001 | [3] |
| ILMN_1807493 | *ACVRL1* | 0.007 | 0.000 | [4] |
| ILMN_1812616 | *MYO1C* | 0.007 | 0.004 | [5] |
| ILMN_2149226 | *CAV1* | 0.007 | 0.009 | [6] |
| ILMN_2410523 | *DDR2* | 0.006 | 0.002 | [7] |
| ILMN_1669772 | *LRP1* | 0.005 | 0.007 | [8, 9, 10] |
| ILMN_1672596 | *BCAR1* | 0.0057 | 0.005 | [11] |
| ILMN_1733811 | *JUP* | 0.007 | 0.010 | [12] |
| ILMN_1743445 | *FAM107A* | 0.0059 | 0.008 | [13] |

Supplementary Table 5 Candidate genes positively associated with 25-OHD level in the GO term ‘*regulation of programmed cell death’* and published evidence of tumour suppressor/ biomarker activity

22 genes from the candidate gene-set were associated with GO term regulation of cell migration, with a total of 162 genes associated with this term.

| ILMN ID | Gene | Coefficient | P value | Reference |
| --- | --- | --- | --- | --- |
| ILMN_1651788 | *MAP3K11* | 0.004 | 0.010 | [14] |
| ILMN_1659058 | *PPP1R10* | 0.004 | 0.004 | [15] |
| ILMN_1685005 | *TNFRSF1A* | 0.004 | 0.002 | [16] |
| ILMN_1687440 | *HIPK2* | 0.004 | 0.006 | [17] |
| ILMN_1721283 | *HSPB6* | 0.008 | 0.001 | [18] |
| ILMN_1669772 | *LRP1* | 0.005 | 0.007 | [8, 9, 10] |
| ILMN_1672596 | *BCAR1* | 0.006 | 0.005 | [11] |
| ILMN_1791447 | *CXCL12* | 0.006 | 0.009 | [19] |
| ILMN_1805990 | *BAK1* | 0.004 | 0.009 | [20] |

Supplementary Table 6 Association between relevant genetic variants and 25OHD fold-change after supplementation

Estimate relates to difference in fold-change in 25-OHD with supplementation per reference allele. E.g. For rs2282679, average fold-change is 3.66, 2.43 and 1.93 with 0, 1, and 2 G alleles respectively.

| Locus | Reference allele | Reference allele frequency | 12 weeks | |
| --- | --- | --- | --- | --- |
|  |  |  | **Estimate** | **P value** |
| rs2282679 | G | 0.29 | -0.73 | 0.03 |
| rs10741657 | A | 0.33 | 0.40 | 0.11 |
| rs2228570 | A | 0.34 | 0.57 | 0.06 |
| rs6013897 | A | 0.14 | 0.13 | 0.76 |

Supplementary Table 7 Direction and magnitude of effect in candidate genes with expression change after supplementation

|  |  | PHASE 1 | | PHASE 2 | |
| --- | --- | --- | --- | --- | --- |
| ID | **Gene** | **Coefficient** | **P** | **Fold-change** | **P** |
| ILMN_1661650 | *SMEK2* | -0.002 | 0.008 | 0.93 | 0.020 |
| ILMN_1666050 | *TMUB1* | 0.005 | 0.005 | 1.14 | 0.012 |
| ILMN_1668345 | *OAF* | 0.005 | 0.009 | 1.10 | 0.039 |
| ILMN_1668639 | *TBC1D10B* | 0.004 | 0.010 | 1.11 | 0.042 |
| ILMN_1670456 | *RBM42* | 0.004 | 0.003 | 1.12 | 0.024 |
| ILMN_1675331 | *PEG3* | 0.002 | 0.001 | 1.06 | 0.046 |
| ILMN_1684929 | *TOPBP1* | -0.004 | 0.008 | 0.91 | 0.023 |
| ILMN_1687440 | *HIPK2* | 0.004 | 0.003 | 1.17 | 0.016 |
| ILMN_1690241 | *BATF2* | 0.004 | 0.006 | 1.11 | 0.036 |
| ILMN_1694325 | *NFIX* | 0.007 | 0.006 | 1.15 | 0.038 |
| ILMN_1701855 | *PPP1CC* | -0.002 | 0.007 | 0.94 | 0.020 |
| ILMN_1710514 | *BCL3* | 0.005 | 0.009 | 1.17 | 0.015 |
| ILMN_1716056 | *LMF2* | 0.004 | 0.007 | 1.14 | 0.010 |
| ILMN_1719656 | *MRPL38* | 0.003 | 0.004 | 1.11 | 0.010 |
| ILMN_1721022 | *SHC1* | 0.002 | 0.009 | 1.07 | 0.026 |
| ILMN_1722798 | *PLCD3* | 0.006 | 0.004 | 1.18 | 0.010 |
| ILMN_1723684 | *DARC* | 0.007 | 0.010 | 1.29 | 0.001 |
| ILMN_1727300 | *ZNF444* | 0.004 | 0.009 | 1.14 | 0.003 |
| ILMN_1727389 | *CDC16* | -0.003 | 0.002 | 0.93 | 0.020 |
| ILMN_1728660 | *PMPCB* | -0.002 | 0.005 | 0.92 | 0.003 |
| ILMN_1732799 | *CD34* | 0.007 | 0.004 | 1.24 | 0.003 |
| ILMN_1734445 | *LOC91461* | 0.006 | 0.005 | 1.17 | 0.034 |
| ILMN_1738116 | *TMEM119* | 0.006 | 0.005 | 1.19 | 0.005 |
| ILMN_1739640 | *DCHS1* | 0.004 | 0.001 | 1.11 | 0.023 |
| ILMN_1739659 | *ZDHHC6* | -0.003 | 0.001 | 0.90 | 0.005 |
| ILMN_1743538 | *MLLT10* | -0.004 | 0.006 | 0.91 | 0.013 |
| ILMN_1747460 | *TMEM184B* | 0.004 | 0.009 | 1.11 | 0.029 |
| ILMN_1751171 | *IRF2BP1* | 0.005 | 0.003 | 1.24 | 0.000 |
| ILMN_1757877 | *HCFC1R1* | 0.004 | 0.007 | 1.10 | 0.041 |
| ILMN_1766798 | *CENTB2* | -0.004 | 0.003 | 0.90 | 0.012 |
| ILMN_1768282 | *SNX21* | 0.004 | 0.003 | 1.08 | 0.047 |
| ILMN_1771120 | *TMEM45B* | 0.007 | 0.007 | 1.34 | 0.005 |
| ILMN_1780357 | *PRRT1* | 0.003 | 0.001 | 1.08 | 0.022 |
| ILMN_1796089 | *NKX2-3* | 0.005 | 0.001 | 1.19 | 0.001 |
| ILMN_1807704 | *LOC338799* | -0.002 | 0.005 | 0.95 | 0.002 |
| ILMN_1811650 | *DUS2L* | -0.003 | 0.008 | 0.91 | 0.012 |
| ILMN_1812262 | *DDR1* | 0.007 | 0.008 | 1.14 | 0.001 |
| ILMN_1813423 | *NAT15* | 0.003 | 0.002 | 1.07 | 0.046 |
| ILMN_1813641 | *C9orf167* | 0.006 | 0.009 | 1.16 | 0.049 |
| ILMN_1814165 | *SSBP3* | 0.004 | 0.007 | 1.13 | 0.029 |
| ILMN_1815937 | *TMEM184A* | 0.004 | 0.006 | 1.19 | 0.005 |
| ILMN_1826130 | *NA* | -0.001 | 0.001 | 0.96 | 0.030 |
| ILMN_2093500 | *ZBED5* | -0.002 | 0.008 | 0.92 | 0.007 |
| ILMN_2113535 | *PCYOX1* | 0.004 | 0.009 | 1.22 | 0.003 |
| ILMN_2251978 | *C21orf66* | -0.003 | 0.004 | 0.91 | 0.043 |
| ILMN_2339796 | *CDC16* | -0.002 | 0.005 | 0.93 | 0.017 |
| ILMN_2362681 | *CES2* | 0.007 | 0.006 | 1.34 | 0.001 |
| ILMN_3242462 | *UHRF1BP1* | -0.004 | 0.008 | 0.92 | 0.022 |
| ILMN_3290428 | *LOC284297* | 0.003 | 0.001 | 1.07 | 0.037 |

**Supplementary Figure 1** **Gene-set enrichment plot for enrichment of candidate genes associated with 25-OHD level in those undergoing vitamin D supplementation**

Enrichment plot y axis charts relative enrichment. Each black bar within the bar code represents a different candidate gene which are ranked from left to right by increasing T statistic from a moderated T test. The curve (or worm) above the barcode shows the relative local enrichment of the bars in each part of the plot. The dotted horizontal line indicates neutral enrichment; the worm above the dotted line shows enrichment while the worm below the dotted line shows depletion. Top row is all participants after supplementation. Second row shows enrichment in participants with <1.5X increase in 25-OHD (average FC=1.26), and third row shows enrichment in participants with >1.5X increase in 25-OHD (average FC=3.37). Bottom row shows enrichment in subset who underwent interval sampling prior to supplementation (i.e. natural state). All plots calculated for 349 genes with positive association with 25-OHD and 206 genes with negative association with 25-OHD.

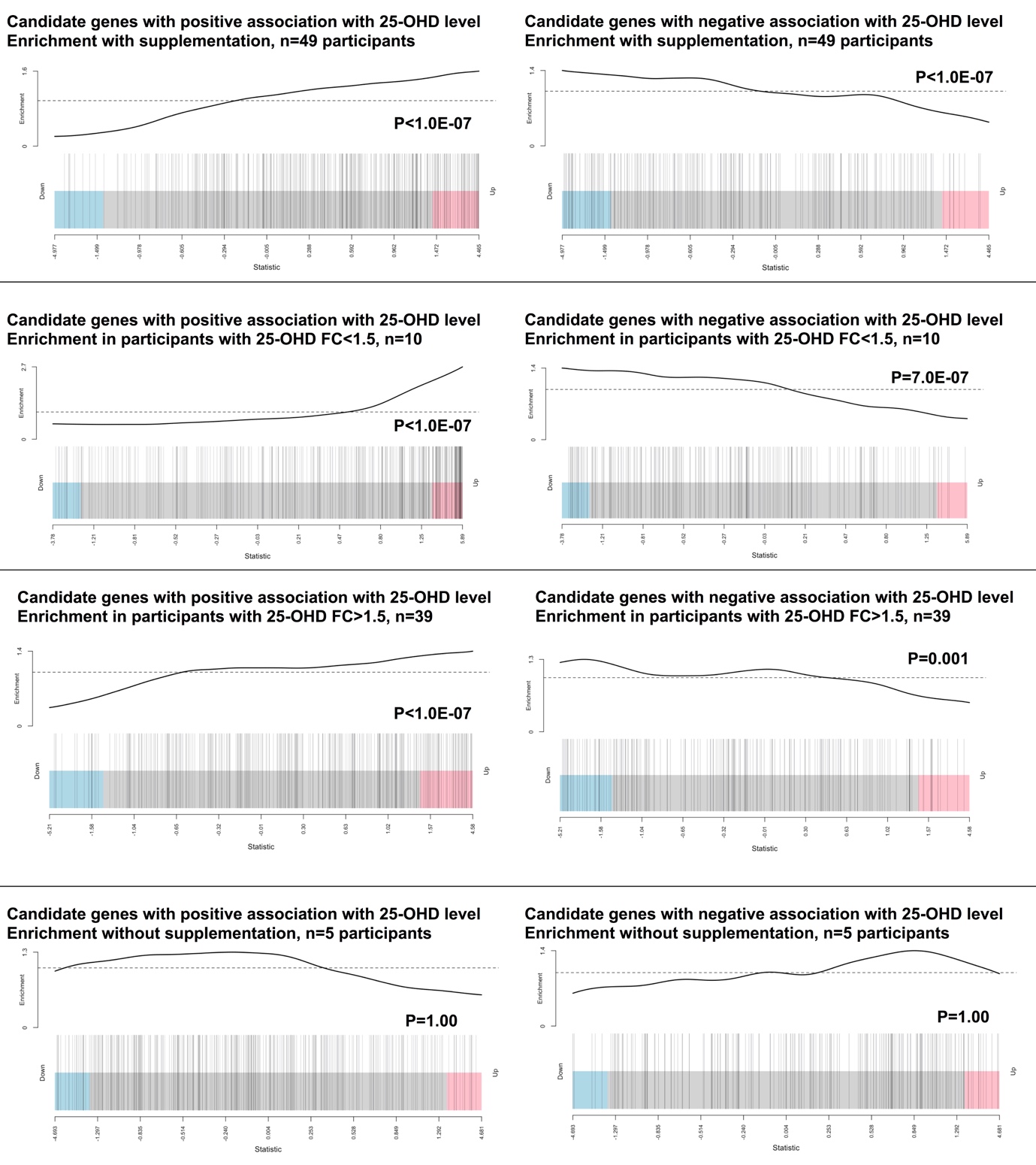

Supplementary Table 8 Top ranked Enriched GO terms after vitamin D supplementation

| GO term | Description | P value | FDR q-value | Enrichment |
| --- | --- | --- | --- | --- |
| GO:0030335 | Positive regulation of cell migration | 1.60E-07 | 2.43E-03 | 2.11 |
| GO:0044238 | Primary metabolic process | 5.61E-07 | 4.24E-03 | 1.15 |
| GO:0010564 | Regulation of cell cycle process | 5.87E-07 | 2.96E-03 | 1.62 |
| GO:2000147 | Positive regulation of cell motility | 7.47E-07 | 2.82E-03 | 2.02 |
| GO:0040017 | Positive regulation of locomotion | 1.15E-06 | 3.48E-03 | 1.98 |
| GO:0051272 | Positive regulation of cellular component movement | 2.55E-06 | 6.43E-03 | 1.96 |
| GO:0090068 | Positive regulation of cell cycle process | 3.23E-06 | 6.98E-03 | 2.21 |
| GO:0045787 | Positive regulation of cell cycle | 3.26E-06 | 6.16E-03 | 1.86 |
| GO:0007346 | Regulation of mitotic cell cycle | 4.01E-06 | 6.73E-03 | 1.75 |
| GO:0045931 | Positive regulation of mitotic cell cycle | 4.23E-06 | 6.39E-03 | 2.56 |
| GO:0038065 | Collagen-activated signalling pathway | 4.46E-06 | 6.13E-03 | 6.83 |
| GO:0038063 | Collagen-activated tyrosine kinase receptor signalling pathway | 4.60E-06 | 5.80E-03 | 7.73 |
| GO:0044237 | Cellular metabolic process | 5.61E-06 | 6.52E-03 | 1.14 |
| GO:1901576 | Organic substance biosynthetic process | 5.77E-06 | 6.23E-03 | 1.3 |
| GO:0009058 | Biosynthetic process | 6.84E-06 | 6.89E-03 | 1.29 |
| GO:0006807 | Nitrogen compound metabolic process | 7.15E-06 | 6.76E-03 | 1.14 |
| GO:0071704 | Organic substance metabolic process | 9.17E-06 | 8.15E-03 | 1.13 |
| GO:0030334 | Regulation of cell migration | 9.32E-06 | 7.83E-03 | 1.62 |
| GO:0046777 | Protein autophosphorylation | 1.22E-05 | 9.71E-03 | 2.24 |
| GO:0043067 | Regulation of programmed cell death | 1.28E-05 | 9.66E-03 | 1.38 |

**Supplementary Figure 2 GO terms enriched in PHASE 1 and PHASE 2**

For each group, genes were ranked by ascending magnitude of coefficient or logFC and entered into the GO enrichment software GOrilla. FDR q-value' is the correction of the above p-value for multiple testing using the Benjamini and Hochberg method [21]. For the i^th^ term (ranked according to p-value) the FDR q-value is (p-value * number of GO terms) / i. Enrichment = (the number of genes in the intersection/ number of genes in the top of the input list)/(total number of genes associated with a specific GO term/ total number of genes). Terms excluded where FDR>0.05

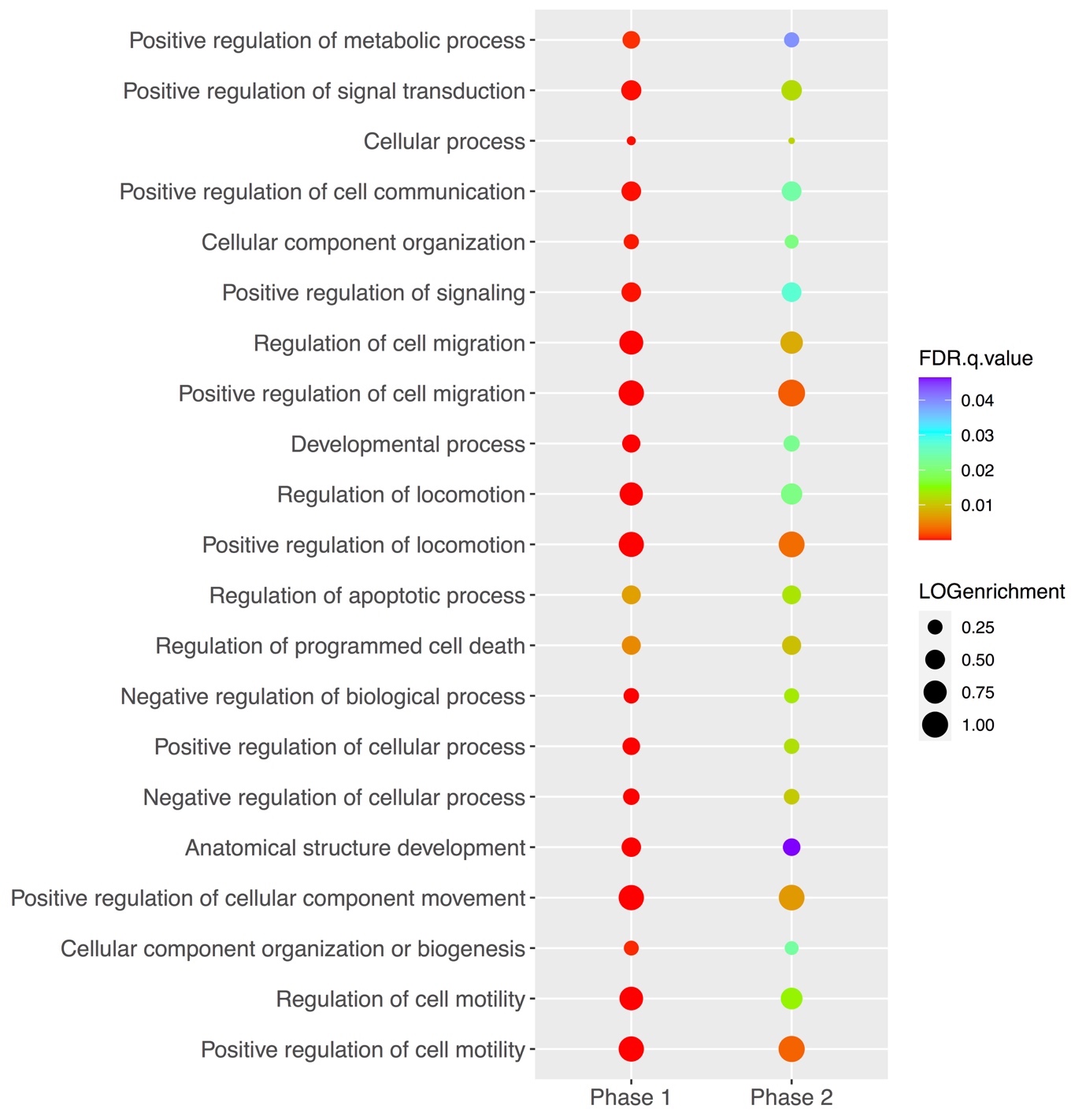

**Supplementary Table 9 Enrichment of candidate GO terms after supplementation in RNAseq data**

Participants with rectal response defined as those with significant enrichment in candidate gene-set from PHASE 1 after supplementation. FDR pvalue given

| GO term | GO term  N genes | N genes tested | All participants | Participants with rectal response | Participants without rectal response |
| --- | --- | --- | --- | --- | --- |
| Top 50 terms | 15142 | 8095 | 2.90E-02 | 1.75E-05 | 0.52 |
| GO:0040011 | 2176 | 1186 | 0.13 | 2.82E-22 | 1.00 |
| GO:0016477 | 1876 | 1047 | 2.54E-02 | 1.00E-24 | 1.00 |
| GO:0009653 | 3385 | 1906 | 0.28 | 1.08E-34 | 1.00 |
| GO:0048870 | 2040 | 1118 | 4.88E-02 | 5.07E-23 | 1.00 |
| GO:0043062 | 455 | 278 | 2.75E-10 | 1.87E-30 | 1.00 |
| GO:0065008 | 5036 | 2827 | 0.45 | 3.06E-03 | 1.00 |
| GO:0022603 | 1444 | 805 | 0.22 | 1.83E-17 | 1.00 |
| GO:0030198 | 455 | 278 | 2.75E-10 | 1.87E-30 | 1.00 |
| GO:0006928 | 2610 | 1434 | 0.89 | 7.63E-19 | 1.00 |
| GO:0032502 | 7761 | 4212 | 0.13 | 3.65E-26 | 1.00 |
| GO:0006936 | 447 | 239 | 1.00 | 2.45E-08 | 1.00 |
| GO:0050793 | 3357 | 1837 | 1.69E-02 | 1.87E-30 | 1.00 |
| GO:0048646 | 1404 | 785 | 6.77E-03 | 1.71E-28 | 1.00 |
| GO:0048856 | 7128 | 3883 | 2.90E-02 | 1.90E-32 | 1.00 |
| GO:0051128 | 3219 | 1756 | 0.90 | 1.62E-03 | 1.00 |
| GO:0003012 | 570 | 309 | 1.00 | 5.08E-08 | 1.00 |
| GO:0022610 | 1816 | 1037 | 5.82E-08 | 7.35E-45 | 1.00 |
| GO:0097435 | 923 | 508 | 1.00 | 2.54E-05 | 1.00 |
| GO:0007155 | 1809 | 1032 | 5.55E-08 | 7.35E-45 | 1.00 |
| GO:0051094 | 1774 | 981 | 1.24E-02 | 3.27E-23 | 1.00 |
| GO:0010273 | 15 | 11 | 1.00 | 1.00 | 1.00 |
| GO:0051239 | 3968 | 2189 | 1.04E-02 | 4.22E-32 | 1.00 |
| GO:0048518 | 7840 | 4333 | 1.15E-02 | 7.38E-13 | 1.00 |
| GO:0030334 | 1166 | 643 | 0.21 | 2.34E-16 | 1.00 |
| GO:2000145 | 1238 | 680 | 0.22 | 1.64E-15 | 1.00 |
| GO:0061687 | 17 | 12 | 1.00 | 1.00 | 1.00 |
| GO:0046688 | 53 | 28 | 0.90 | 0.95 | 1.00 |
| GO:0048583 | 5400 | 3008 | 6.14E-06 | 2.40E-19 | 1.00 |
| GO:0051270 | 1351 | 738 | 0.47 | 2.54E-15 | 1.00 |
| GO:0043408 | 979 | 540 | 0.13 | 4.74E-09 | 1.00 |
| GO:0048522 | 7180 | 3970 | 2.04E-02 | 3.10E-12 | 1.00 |
| GO:0040012 | 1289 | 707 | 0.31 | 4.91E-15 | 1.00 |
| GO:0040008 | 888 | 489 | 0.53 | 2.29E-06 | 1.00 |
| GO:0007166 | 3793 | 2139 | 1.36E-08 | 2.30E-29 | 1.00 |
| GO:0048523 | 6410 | 3432 | 0.58 | 6.76E-09 | 1.00 |
| GO:0010035 | 742 | 421 | 0.78 | 0.85 | 1.00 |
| GO:0045597 | 1266 | 713 | 0.09 | 4.27E-18 | 1.00 |
| GO:0098754 | 184 | 97 | 0.40 | 1.00 | 0.005 |
| GO:0048519 | 7396 | 3849 | 0.44 | 1.20E-04 | 1.00 |
| GO:0023051 | 4691 | 2616 | 2.14E-03 | 3.97E-21 | 1.00 |
| GO:0009966 | 4105 | 2282 | 2.69E-04 | 4.27E-18 | 1.00 |
| GO:0051272 | 760 | 405 | 0.08 | 2.53E-11 | 1.00 |
| GO:0030029 | 1039 | 580 | 1.00 | 1.25E-10 | 1.00 |
| GO:0006882 | 47 | 28 | 0.77 | 1.00 | 1.00 |
| GO:0048585 | 2173 | 1227 | 2.14E-03 | 2.54E-09 | 1.00 |
| GO:0042127 | 2117 | 1153 | 2.96E-04 | 1.85E-17 | 1.00 |
| GO:0042221 | 5632 | 3099 | 0.13 | 9.45E-05 | 1.00 |
| GO:2000026 | 2653 | 1450 | 4.13E-03 | 1.17E-30 | 1.00 |
| GO:0040017 | 751 | 404 | 0.10 | 1.23E-10 | 1.00 |
| GO:0009887 | 1279 | 746 | 2.14E-03 | 1.10E-22 | 1.00 |

**Supplementary Table 10 Characteristics of participants with rectal mucosa response to supplementation**

Results from multivariate logistic regression model including all factors listed. Baseline 25-OHD is not associated with ‘response’ when instead of 25-OHD fold-change (estimate 0.04, P=0.08).

|  | Participants with rectal mucosa response | Participants without rectal mucosa response | Estimate | p |
| --- | --- | --- | --- | --- |
| Patients | 9 | 40 | - | - |
| Age (median) | 69 | 58 | 0.12 | 0.07 |
| Gender (F) | 4 (45%) | 50% | 0.38 | 0.78 |
| BMI (mean) | 27.4 | 28.6 | 0.09 | 0.40 |
| Past history of CRC | 4 (45%) | 17 (43%) | 0.93 | 0.44 |
| VDR SNP allele score | 4 | 3 | 2.22 | 0.006 |
| *VDR* Fold-change >1 | 5 (55%) | 16 (40%) | 2.77 | 0.11 |
| 25-OHD fold-change at 12 weeks* | 1.6 | 2.8 | -1.96 | 0.02 |

**Supplementary Table 11 AUC values for blood biomarkers of rectal and blood response in the SCOVIDS and BEST-D studies**

C-statistic (AUC) values for predictive utility of blood expression change in blood biomarkers in identifying participants with rectal mucosa or blood expression response to supplementation. Response based on candidate gene-set from PHASE 1 of SCOVID study after HIPK2, PPP1CC, SMEK2, DDR1, SNX21 were excluded. Thresholds used were those derived as best cut-off for predicting rectal mucosa response.

|  | SCOVIDS Rectal response | SCOVIDS Blood response | BEST-D Blood response |
| --- | --- | --- | --- |
|  | AUC (95%CI) | AUC (95%CI) | AUC (95%CI) |
| *SMEK2* FC < 0.68 | 0.72 (0.55-0.89) | 0.55 (0.43-0.71) | NA |
| *HIPK2* FC > 1.19 | 0.75 (0.59-0.87) | 0.77 (0.63-0.88) | 0.74 (0.66-0.81) |
| *PPP1CC* FC < 1.19 | 0.80 (0.63-0.94) | 0.83 (0.67-0.96) | 0.83 (0.76-0.89) |
| *SNX21* FC > 0.88 | 0.72 (0.63-0.81) | 0.59 (0.42-0.74) | 0.53 (0.51-0.56) |
| *DDR1* <>0.95 | 0.65 (0.46-0.82) | 0.59 (0.42-0.74) | 0.44 (0.49-0.62) |
| *All genes* | 0.99 (0.97-1.00) | 0.91 (0.80-1.00) | 0.89 (0.83-0.95) |
| *HIPK2* + *PPP1CC* | 0.84 (0.66-1.00) | 0.87 (0.71-1.00) | 0.85 (0.78-0.91) |

**Supplementary Figure 3 Receiver-operator curve for putative blood biomarkers and rectal mucosal response to supplementation**

ROC curve of predictive utility in identifying participants with rectal mucosa response to supplementation based on blood expression change with supplementation of SMEK2 FC< 0.68 + HIPK2 FC> 1.19 + PPP1CC FC< 1.19 + SNX21 FC >0.88 + DDR1<>0.95

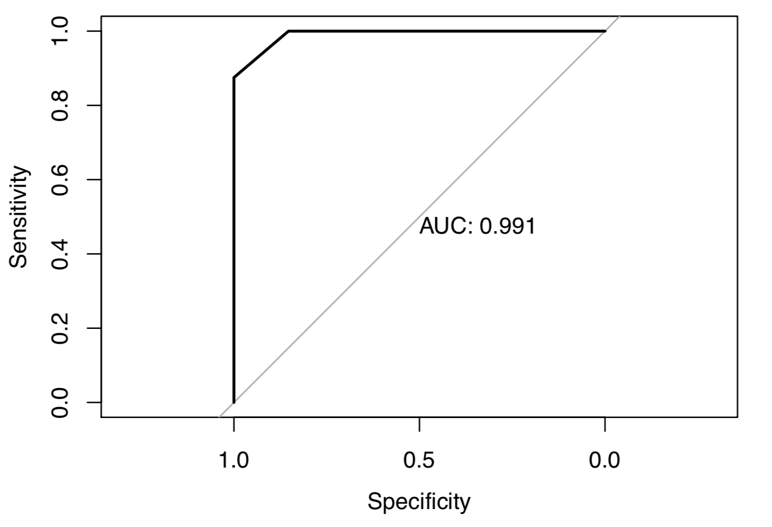
