## Supplementary Methods for "Oral Vitamin D supplementation induces transcriptomic changes in rectal mucosa that are consistent with anti-tumour effects"

**Peripheral blood mononuclear cell extraction**

Blood was sampled by standard venepuncture of a peripheral arm vein, with plasma and PBMCs extracted using Ficoll-Paque Plus (GE Healthcare Life Sciences, UK). The sampled blood was layered over an equal volume of Ficoll-Paque and spun at 2,500 rpm for 15 minutes. The upper layer (plasma) was taken off and frozen at -40°C. The lymphocyte layer was then transferred to a clean centrifuge tube and spun at 2,500 rpm followed by a wash in ice-cold PBS. The lymphocyte layer was then spun down and the pellet stored at -80°C or immediately immersed in TRIzol (Applied Biosystems) for downstream RNA extraction.

**PBMC and mucosa sample processing for gene expression analysis**

RNA was extracted from PBMCs and mucosa using a proprietary RNA extraction kit (Ribopure kit, Applied Biosystems) according to the manufacturer’s protocol. This process initially involved homogenising the sample in 1 mL TRIzol. The homogenates were then incubated for 5 minutes at room temperature to enable the nucleoprotein complexes to completely dissociate. 200 μl of chloroform was added and the tubes vortexed at 2800 rpm for 15 seconds. After 5 minutes’ incubation at room temperature, the homogenate was centrifuged at 12,000 x g for 10 min at 4°C. This separated the homogenate into a lower, red, organic phase (phenol-BCP phase); an interphase; and a colourless, upper, aqueous phase. 400 μl of the aqueous phase was withdrawn to which 200 μl of ethanol was added. The mixture was vortexed at maximum speed for 5 seconds and added to a filter cartridge-collection tube. This was centrifuged at 12,000 x g for 30 seconds at room temperature and the flow-through discarded. Two wash cycles using 500 μl of provided wash solution were undertaken, with a final spin of 12,000 x g for 30 seconds to ensure all wash solution had left the filter. Finally, 50 μl of provided elution buffer was added to the filter cartridge within a new collection tube and spun at 12,000 x g for 30 seconds to elute the RNA. RNA purity (A260/A280 and A260/A230) and concentration was assessed by the Nanodrop 800 spectrophotometer. The RNA yield and integrity of samples to be submitted for microarray expression analysis was assessed using the 2100 Bioanalyzer®.

**Assessment of rectal mucosa gene expression**

For HT12 analysis, total RNA was converted to double-stranded cDNA, followed by in vitro transcription amplification to generate labelled cRNA (Illumina TotalPrep™ RNA Amplification Kit). Gene expression profiling was undertaken using the HumanHT-12 v4.0 Expression BeadChip Arrays (Illumina) and IScan NO660 scanner, providing coverage of 47,231 transcripts and >31,000 annotated genes derived from the National Centre for Biotechnology Information Reference Sequence RefSeq Release 38 (November 7, 2009). Microarray data was exported from BeadStudio (Illumina) and processed in R using the *limma* package. In brief, the steps involved were background correction using negative controls, quantile normalization to remove technical variation and finally log2 transformation. Control probes and probes that were not expressed in at least three arrays to a detection-value of ≥5% were excluded. A standard approach to batch correction using ComBat was performed to control for batch effects [1].

**RNA sequencing and analysis**

Whole-genome [transcriptomic](https://www.sciencedirect.com/topics/medicine-and-dentistry/transcriptomics) patterns were analysed on total RNA from selected normal mucosa samples. RNA samples from the human supplementation study (0 and 12 week samples) were subjected to 150bp paired-end total RNA-seq (155M reads) in a single batch. RNA integrity and yield was quantified using the 2100 Bioanalyzer®. Extracted RNA was reverse transcribed using Moloney Murine Leukemia Virus reverse transcriptase and random primers (Promega) at 37°C for 30 minutes and 95°C for 5 minutes. RNA samples were submitted to the Edinburgh Genomics sequencing facility, where QC, ribosomal-depletion, strand-aware library preparation and Illumina adapter ligation was performed. Ribosomal RNA was depleted using the New England Biolabs NEBNext rRNA Depletion Kit according to the manufacturer’s protocol. Samples were sequenced on the Illumina HiSeq 2500 platform in “rapid mode” with 150bp paired-end reads. Transcript indexing and quantification from RNA-seq reads was performed using Salmon v1.1.0[2]. The Salmon index was generated using the reference dataset GRCh38.primary_assembly.fa and the Ensembl 96 release, (<ftp://ftp.ensembl.org/pub/release-96/fasta/homo_sapiens)>. In brief, the Ensembl (cDNA and ncRNA) and reference FASTA were concatenated, the ‘decoys.txt’ was prepared from GRCh38 and the Salmon index run on the concatenated FASTA file in the R statistical environment (v4.0.0). Paired reads 1 and read 2 fastq files respectively were then concatenated for each sample, and Salmon run using auto-detect strandedness and ‘validateMappings’ flag.

Salmon transcript level quantitation files were combined and summarised to gene-level counts using txiimport[3] and biomaRt[4, 5]. Transcript level counts were aggregated into gene levels counts and imported into R using the tximport package. A Differential Gene Expression object (DGEList) was created using tximport generated gene-level counts and sample metadata for analysis using edgeR (3.30.0) and limma (3.44.1). Genes were filtered using the filterByExpr function in the edgeR package (3.30.0) to include genes with at least 10 reads per gene. The calcNormFactors function was used to calculate Trimmed mean of M-values (TMM) between-sample normalisation to scale the library size. The limma ‘voom’ function was used to transform normalized count data to log2-counts per million (logCPM), estimate the mean-variance relationship and used to compute appropriate observation-level weights.

**Genotype at the vitamin D pathway loci**

Blood leukocyte DNA was genotyped for four polymorphisms known to be associated with 25-OHD level [6]: rs2282679, in *GC;* rs12785878, near *DHCR7*; rs10741657, near *CYP2R1*; rs6013897 near *CYP24A1* to test for association with 25-OHD change after vitamin D supplementation. Four polymorphisms in the *VDR* gene which are established as functionally relevant, impacting *VDR* function, DNA binding and calcitriol binding (rs1544410, rs10735810, rs7975232, rs11568820) were also genotyped by using either an Illumina Infinium array or obtained from whole-genome sequencing data. A *VDR* allele score was derived by summing the following allele counts from four functionally relevant SNPs in the vitamin D receptor gene: rs10735810 A allele, a variant that determines the translational start site and downstream 1,25-dihydroxycholecalciferol effects [7], rs1544410 T allele which has been associated with increased *VDR* mRNA expression[8], rs11568820 T allele, a variant that is located in the *VDR* promoter region and directly influences transcriptional activity [9], and

rs7975232 A allele which impacts mRNA stability [7].

**Statistical analysis**

Receiver operator curves and C statistic were derived using the ‘prediction’ and ‘performance’ functions within the ‘pROC’ package in R [10]. AUC confidence intervals were calculated using a bootstrap method (x10,000). The best threshold of gene expression fold-change for predicting response was calculated using the ‘coords’ function within the ‘pROC’ package. We then assessed the performance of the same biomarkers in identifying those with ‘response’ to the candidate gene-set in the blood, as a surrogate marker for response in NM.

10	R Development Core Team. R: A language and environment for statistical computing. R Foundation for Statistical Computing, Vienna, Austria, 2013.
